# The Use of Machine Learning Methods in Neurodegenerative Disease Research: A Scoping Review

**DOI:** 10.1101/2023.07.31.23293414

**Authors:** Antonio Ciampi, Julie Rouette, Fabio Pellegrini, Gabrielle Simoneau, Bastien Caba, Arie Gafson, Carl de Moor, Shibeshih Belachew

## Abstract

Machine learning (ML) methods are increasingly used in clinical research, but their extent is complex and largely unknown in the field of neurodegenerative diseases (ND). This scoping review describes state-of-the-art ML in ND research using MEDLINE (PubMed), Embase (Ovid), Central (Cochrane), and Institute of Electrical and Electronics Engineers Xplore. Included articles, published between January 1, 2016, and December 31, 2020, used patient data on Alzheimer’s disease, multiple sclerosis, amyotrophic lateral sclerosis, Parkinson’s disease, or Huntington’s disease that employed ML methods during primary analysis. One reviewer screened citations for inclusion; 5 conducted data extraction. For each article, we abstracted the type of ND; publication year; sample size; ML algorithm data type; primary clinical goal (disease diagnosis/prognosis/prediction of treatment effect); and ML method type. Quantitative and qualitative syntheses of the results were conducted. After screening 4,471 citations and searching 1,677 full-text articles, 1,485 articles were included. The number of articles using ML methods in ND research increased from 172 in 2016 to 490 in 2020, with most of those in Alzheimer’s disease. The most common data type was imaging data (46.9% of articles), followed by functional (20.6%), clinical (14.2%), biospecimen (6.2%), genetic (5.9%), electrophysiological (5.1%), and molecular (1.1%). Overall, 68.5% of imaging data studies were in Alzheimer’s disease and 75.9% of functional data studies were in Parkinson’s disease. Disease diagnosis was the most common clinical aim in studies using ML methods (73.5%), followed by disease prognosis (21.4%) and prediction of treatment effect (13.5%). We extracted 2,734 ML methods, with support vector machine (n=651, 23.8%), random forest (n=310, 11.3%), and convolutional neural network (n=166, 6.1%) representing the majority. Finally, we identified 322 unique ML methods. There are opportunities for additional research using ML methods for disease prognosis and prediction of treatment effect. Addressing these utilization gaps will be important in future studies.

**Author Summary:** Few state-of-the-art scientific updates have been targeted for broader readerships without indulging in technical jargon. We have learned a lot from Judea Pearl on how to put things into context and make them clear. In this review paper, we identify machine learning methods used in the realm of neurodegenerative diseases and describe how the use of these methods can be enhanced in neurodegenerative disease research.

## Introduction

One of the foremost economic, social, and medical challenges of our time is the phenomena of an aging population and the associated burden of chronic diseases.^1^ Among these, neurodegenerative diseases are pathological processes causing progressive deterioration of the neurons of the central nervous system (brain and spinal cord), which manifest as progressively disabling symptoms for patients, generally leading to total loss of autonomy and death. The ensuing human distress can be gleaned from measurable epidemiological metrics such as deaths and absolute number of disability-adjusted life-years.^2^ Without major innovations for disease modification, it is estimated that governments will face increasing demands for care of neurodegenerative diseases such as Alzheimer’s disease (AD), Parkinson’s disease (PD), multiple sclerosis (MS), amyotrophic lateral sclerosis (ALS), and Huntington’s disease (HD).

In response to this challenge, a major scientific effort is underway to tackle individualized treatment predictions to support a paradigm shift towards prediction medicine. For this, the focus has mainly been the initial challenges of defining the disease and looking for treatment. The objectives of this effort are to (1) understand the pathological processes underlying neurodegenerative diseases; (2) develop effective diagnostic and prognostic tools; (3) obtain detailed descriptions of clinical courses; and (4) discover and evaluate treatment approaches. The effort proceeds in the context of expanding availability of computer tools for collecting and analyzing data. In parallel, various disciplines including mathematics, statistics, and computer sciences, among others, converge in a relatively new field called data science.^3^ Data science intersects with artificial intelligence (AI), of which machine learning (ML) is a subfield.

Previous reviews have shown that the use of ML in neurodegenerative disease is rapidly increasing.^4–6^ We felt that the situation called for a scoping review, which is defined as a "preliminary assessment of potential size and scope of the available research literature".^7^ Two recent reviews were of high quality,^8, 9^ but did not qualify as scoping reviews because no description of the method used to retrieve the data from the literature was offered. In contrast, 2 recent papers^6, 10^ were designed as scoping reviews, however, both articles reviewed a broad disease scope (neurological and psychiatric diseases), but narrow scopes in terms of data type (magnetic resonance imaging only) and ML methods. Therefore, we felt the need to carry out a scoping review that would focus on ML methods used in the study of the 5 most prevalent neurodegenerative diseases. In this paper, we present the methods and results of such a review.

## Methods

### Protocol

We developed a scoping review protocol using methodological guidance from the Joanna Briggs Institute for the conduct of scoping reviews to establish a population, concept, and context framework.^11^ The protocol was reviewed by the research team and then further revised.

### Eligibility Criteria

We included any articles (1) in the field of AD, MS, ALS, PD, and/or HD; (2) that included patient data, either from observational studies or randomized controlled trials; (3) that used ML methods in their primary analysis; and (4) that were conducted for the clinical goal of disease diagnosis, disease prognosis, or prediction of treatment effect. We included all study designs except ecological studies, non–peer reviewed literature, meta-analyses, and methodological or simulation studies.

### Search Strategy and Sources

A comprehensive search strategy was created through the identification of keywords by medical librarians and informed by previously published reviews in ML methods and/or neurodegenerative diseases.^8–10, 12–14^ A total of 4 electronic databases were searched from January 1, 2016, to December 31, 2020: MEDLINE (PubMed), Embase (Ovid), Central (Cochrane) and Institute of Electrical and Electronics Engineers Xplore. We had intended to include articles from database inception to December 31, 2020, but had to restrict the search period from 2016 to 2020 to optimize feasibility while retaining breadth and comprehensiveness, allowing the identification of articles using contemporary ML methods. We also limited the language to articles published in English owing to the large number of articles identified. The final search strategy for MEDLINE (PubMed) is presented in S1 Method.

### Study **S**election

Search results were imported into EndNote. A screening questionnaire was developed by the research team a priori. Prior to screening, all reviewers (J.R., A.A., S.A., Z.O., K.Z., G.W.) met and discussed the inclusion and exclusion criteria for the scoping review and gave feedback on the screening questionnaire. The screening questionnaire was then modified based on the feedback. Subsequently, all reviewers independently screened a sample of 10 citations (titles and abstracts) to pilot test the screening questionnaire and ensure clarity of the inclusion and exclusion criteria. A high level of agreement (>80%) needed to be reached before moving on to the next step. One pilot testing session was needed to achieve 83% agreement. The screening questionnaire was then further refined after feedback from pilot testing. For screening, 1 independent reviewer (A.A.) screened titles and abstracts while 1 verifier (J.R.) independently reviewed a sample of the citations to ensure appropriate inclusion and exclusion, and addressed any citations that needed reconciliation.

### Data Extraction

A data extraction form was created a priori in collaboration with the research team. Prior to data extraction, all reviewers (J.R., A.A., S.A., Z.O., K.Z., G.W.) discussed the data to be extracted from each full-text article and provided feedback on the data extraction form. The form was then modified based on the feedback. Then, a pilot test of the form was conducted to ensure clarity and accuracy of the extraction, for which all reviewers independently extracted data from a sample of 10 full-text articles. A high level of agreement (>80%) needed to be reached before moving on to the next step. Two pilot sessions for data extraction were conducted to reach a final level of agreement of 90%. Subsequently, 5 reviewers (A.A., S.A., Z.O., K.Z., G.W.) independently extracted data from the full-text articles while 1 verifier (J.R.) independently performed quality checks and addressed full-text articles needing reconciliation. Two verifiers (A.A., J.R.) then inspected the final database of extracted data to ensure accuracy and performed data cleaning to ensure data consistency. The data extracted from the included full-text articles comprised (1) first author name; (2) type of neurodegenerative disease; (3) publication year; (4) study sample size; (5) data type used in the ML algorithm (clinical, electrophysiological, functional, genetic, imaging, molecular, and biospecimen); (6) primary clinical goal of study for which the ML method was used (disease diagnosis, disease prognosis, or prediction of treatment effect); (7) ML algorithm used in the primary analysis; (8) reason for exclusion (if applicable); and (9) publication details (e.g., journal, volume). Consistent with guidance on scoping reviews, we did not conduct an appraisal of the methodological quality of the included articles.^15, 16^

### Quantitative Data Synthesis

We performed a quantitative analysis of the scoping review, which consisted of frequency analyses. We calculated time trends in the number of articles published per year between 2016 and 2020 for AD, PD, MS, ALS, and HD. For each disease, we calculated the frequency and proportion of articles by data type (clinical, electrophysiological, functional, genetic, imaging, molecular, and biospecimen), and by data type dependent on the clinical aim of the study (disease diagnosis, disease prognosis, and prediction of treatment effect). We presented the ML methods extracted from the review and their frequency. ML methods that were classified as “Other” during the data extraction phase were independently reviewed by 3 reviewers (A.A., J.R., A.C.) to verify their eligibility.

### Qualitative Data Synthesis

We conducted a qualitative analysis of the ML methods extracted from the full-text articles, aiming to define overarching themes for the ML methods extracted from the scoping review. Two reviewers (A.C., J.R.) independently conducted the initial categorization (i.e., theme analysis) by identifying and coding each ML method into non–mutually exclusive themes. The final list of ML methods and their assigned themes were approved by the research team. Because of the high number of ML methods identified, a word cloud of the overarching themes (rather than the individual ML methods) was then created.

## Results

The scoping review included a total of 4,471 screened citations (Figure 1). After screening, 2,727 records were excluded, resulting in the final inclusion of 1,485 full-text articles. The complete list is available upon request.

**Figure 1.**
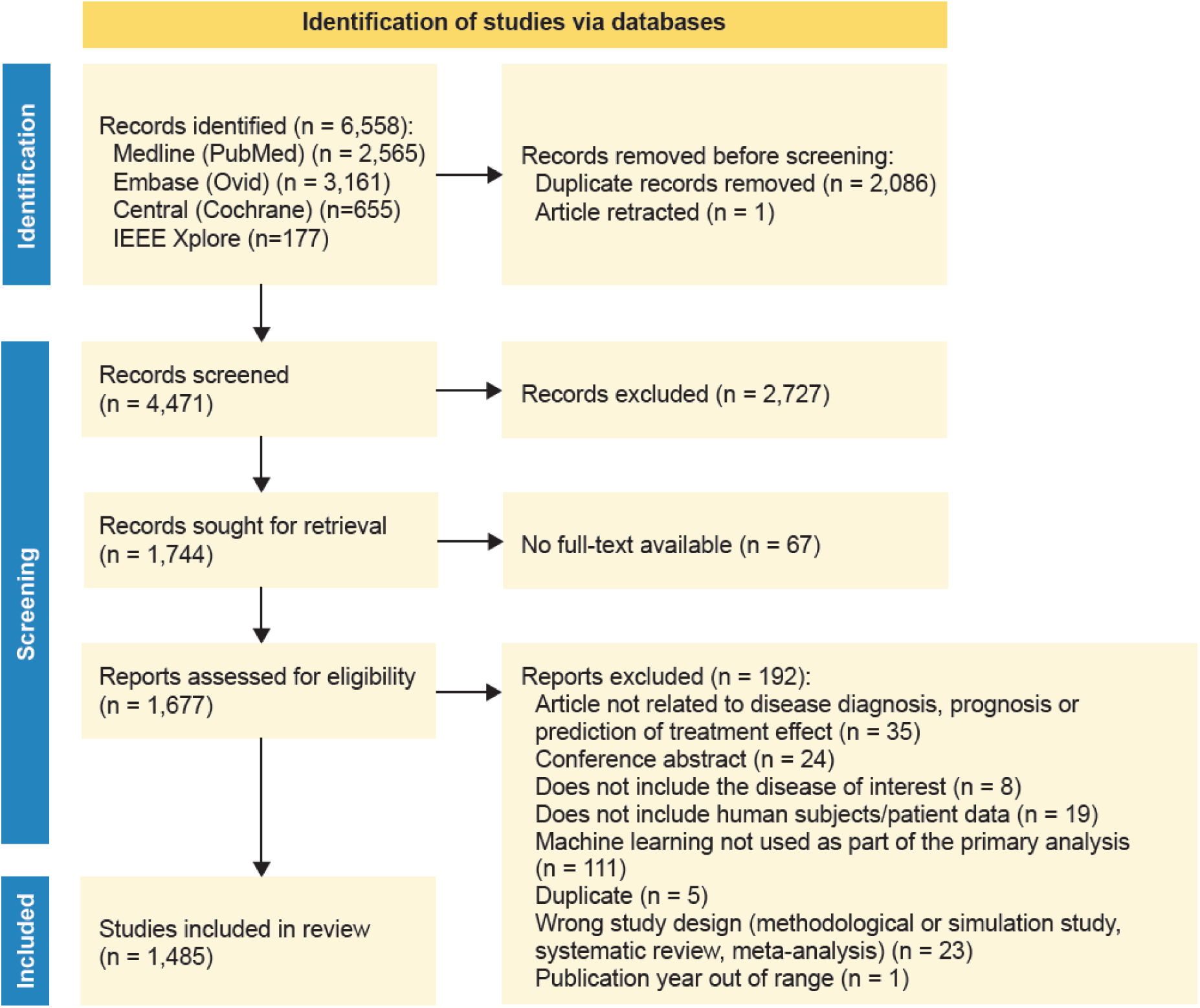
PRISMA Study Flowchart.

Overall, the number of articles using ML methods in neurodegenerative disease increased over time. A total of 172 articles were published in 2016, 217 in 2017, 251 in 2018, 355 in 2019, and 490 in 2020, representing a 185% increase over the total publication period analyzed. AD and PD were the neurodegenerative diseases with the highest number of articles using ML methods, with a total of 776 articles and 497 articles, respectively, compared with 16 articles for HD (Figure 2). The sample size of studies varied widely depending on the disease (S1 Table). Overall, studies included a median of 151 patients (interquartile range, 63–459) and the number of patients was highest for AD (median, 300; interquartile range, 111–737).

**Figure 2.**
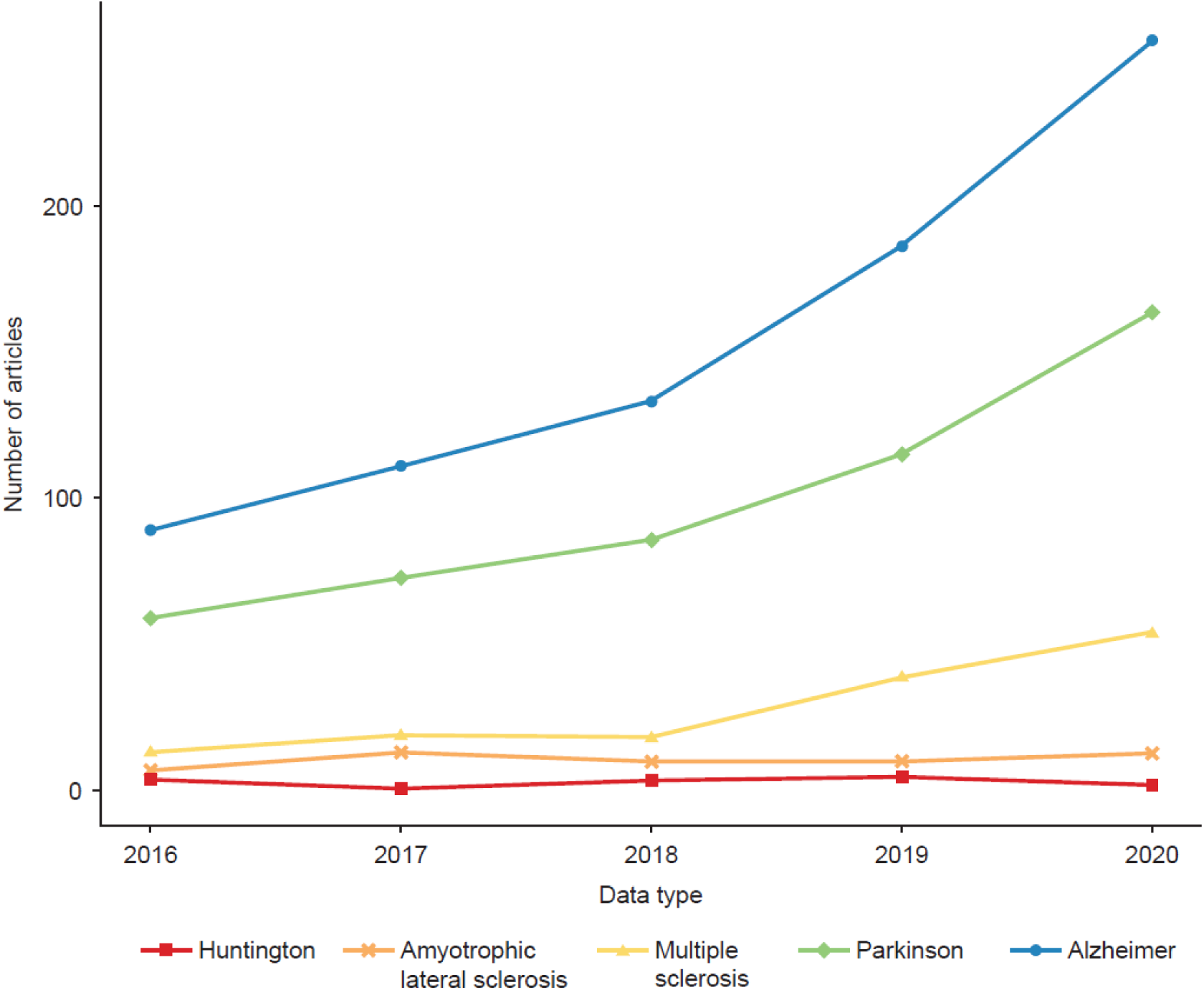
Time Trends in Articles Using Machine Learning Methods in Neurodegenerative Diseases.

The most common data type used in ML was imaging data (n = 813, 46.9% of articles), followed by functional data (n = 357, 20.6%), clinical data (n = 246, 14.2%), biospecimen data (n = 108, 6.2%), genetic data (n = 102, 5.9%), electrophysiological data (n = 89, 5.1%), and molecular data (n = 20, 1.1%) (Figure 3). Figure 4 presents the proportion of studies by data type for each disease, demonstrating which data types are most popular in each disease area. Imaging data were the most commonly used data type in studies in AD (n = 557, 59.3%), MA (n = 91, 56.5%), and ALS (n = 17, 30.4%), whereas functional data represented the most common data type in PD (n = 271, 48.4%) and HD (n = 9, 50.0%).

**Figure 3.**
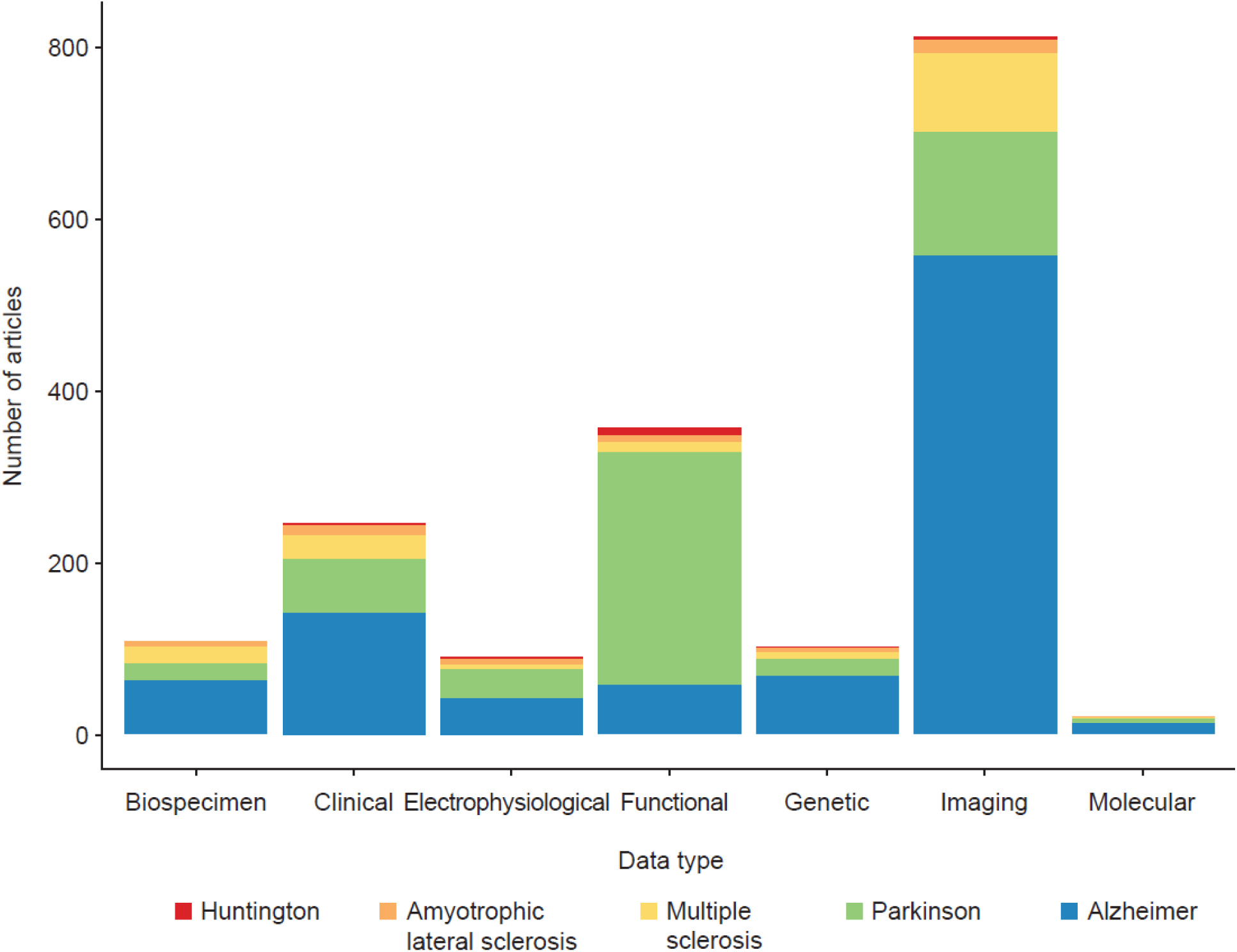
Frequency of Data Type Used in Articles Using Machine Learning Methods, by Neurodegenerative Disease. *Footnote:* An article can include more than 1 data type.

**Figure 4.**
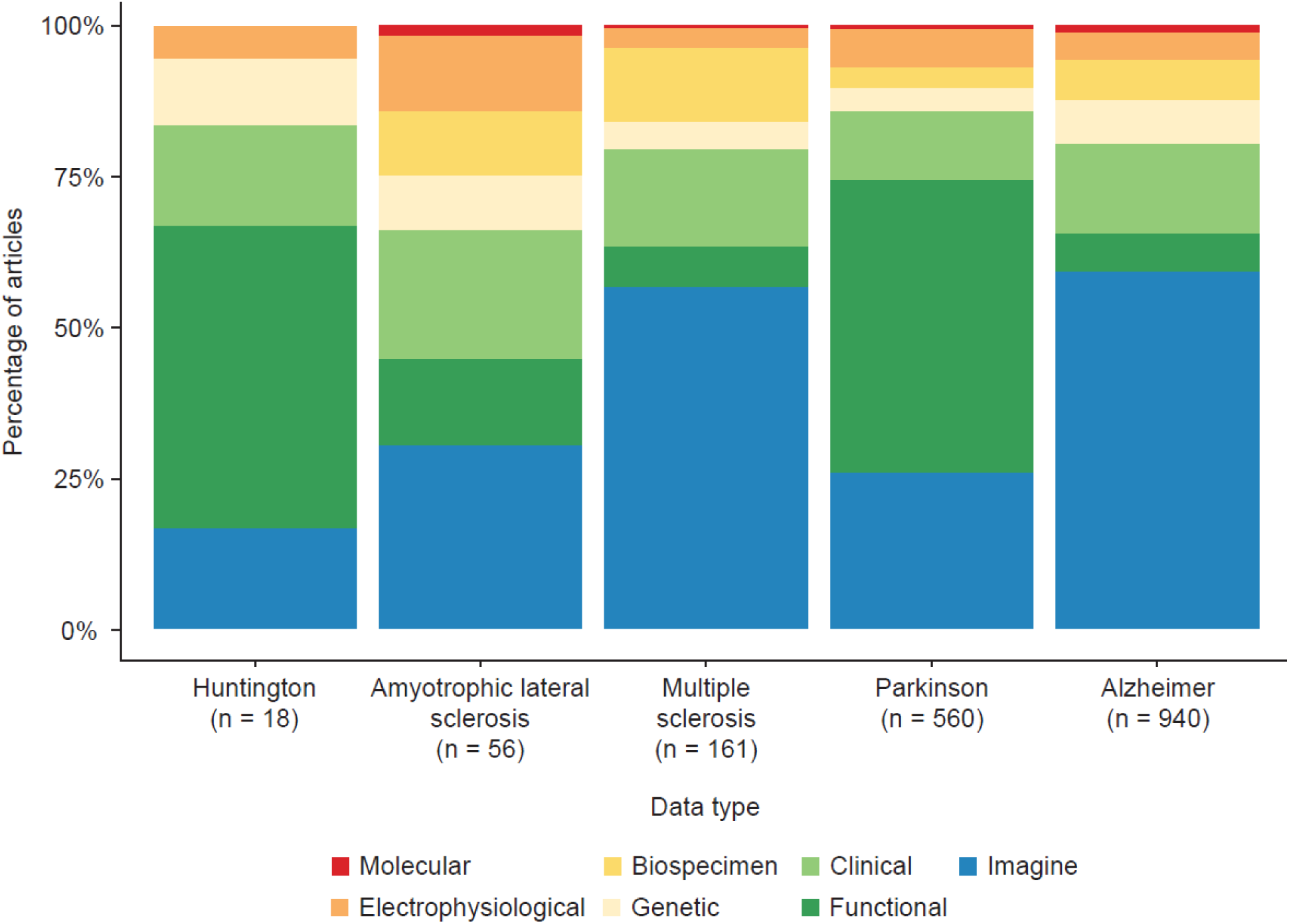
Proportion of Data Type Used in Articles Using Machine Learning Methods, by Neurodegenerative Disease.

Disease diagnosis was the most common clinical objective for studies using ML methods (n = 1,276, 73.5%), followed by disease prognosis (n = 373, 21.4%) and prediction of treatment effect (n = 86, 13.5%). For disease diagnosis, the most common data type was imaging data (n = 636, 49.8%) (S1 Fig.). Studies in AD represented the majority that used all data types except functional data, accounting for 70.7% of imaging data (n = 450), 59% of clinical data (n = 88), 73.6% of genetic data (n = 53), and 59.7% of electrophysiological data (n = 40). Studies in PD represented the majority that used functional data (n = 192, 75.5%). When disease diagnosis was the clinical objective, imaging data, functional data, clinical data, genetic data, and electrophysiological data were used in studies in all 5 neurodegenerative diseases. Biospecimen data were used in all neurodegenerative diseases except HD, and molecular data were only used in AD and PD. Imaging data remained the most common data type for AD, MS, and ALS, whereas functional data represented the most common data type in PD and HD (S2 Fig.).

For disease prognosis, the most common data type was also imaging data (n = 161, 43.1%), followed by clinical data (n = 79, 21.1%) and functional data (n = 72, 19.3%) (S3 Fig.). Studies in AD represented the majority that used imaging data (n = 103, 63.9%), clinical data (n = 48, 60.7%), and biospecimen data (n = 12, 75.0%), whereas studies in PD represented the majority that used functional data (n = 50, 69.4%) and electrophysiological data (n = 12, 75.0%). When disease prognosis was the clinical objective, imaging data, functional data, and clinical data were used in studies in all 5 neurodegenerative diseases. Electrophysiological data, biospecimen data, genetic data, and molecular data were used in few studies (n ≤ 3) in MS, ALS, and HD. Imaging data remained the most common data type in AD and MS studies with disease prognosis as the clinical aim, whereas clinical and functional data were most common in ALS and PD, respectively, and all of imaging, clinical, and functional data were equally used in HD (S4 Fig.).

For prediction of treatment effect, the most common data type was functional data (n = 31, 36.0%) (S5 Fig.). Studies in PD represented all or the majority that used electrophysiological data (n = 6, 100.0%), functional data (n = 29, 93.5%), clinical data (n = 10, 55.5%), and imaging data (n = 7, 43.7%). None of the data types were used in all 5 neurodegenerative diseases. PD was the only disease area using all data types, whereas ALS used clinical data in the single article with treatment effect prediction as a clinical aim (S6 Fig.).

The inclusion of the 1,485 full-text articles resulted in the capture of 2,741 ML methods. Further inspection resulted in the exclusion of 7 ML methods, for a total of 2,734. The total number of full-text articles remained unchanged. The majority of the ML methods extracted were support vector machine (n = 651, 23.8%), random forest (n = 310, 11.3%), and convolutional neural network (n = 166, 6.1%) (S2 Table). We identified a total of 322 unique ML methods in the scoping review. These methods were classified into non–mutually exclusive themes including 8 overarching themes (supervised methods, unsupervised methods, Bayesian methods, ensemble methods, reinforcement learning, neural net, deep learning, and bionet) and 56 subthemes (S3 Table). A word cloud displays the most commonly assigned themes, with supervised methods being the most common (42.0%), followed by unsupervised methods (23.3%) and neural net (17.8%) (Figure 5).

**Figure 5.**
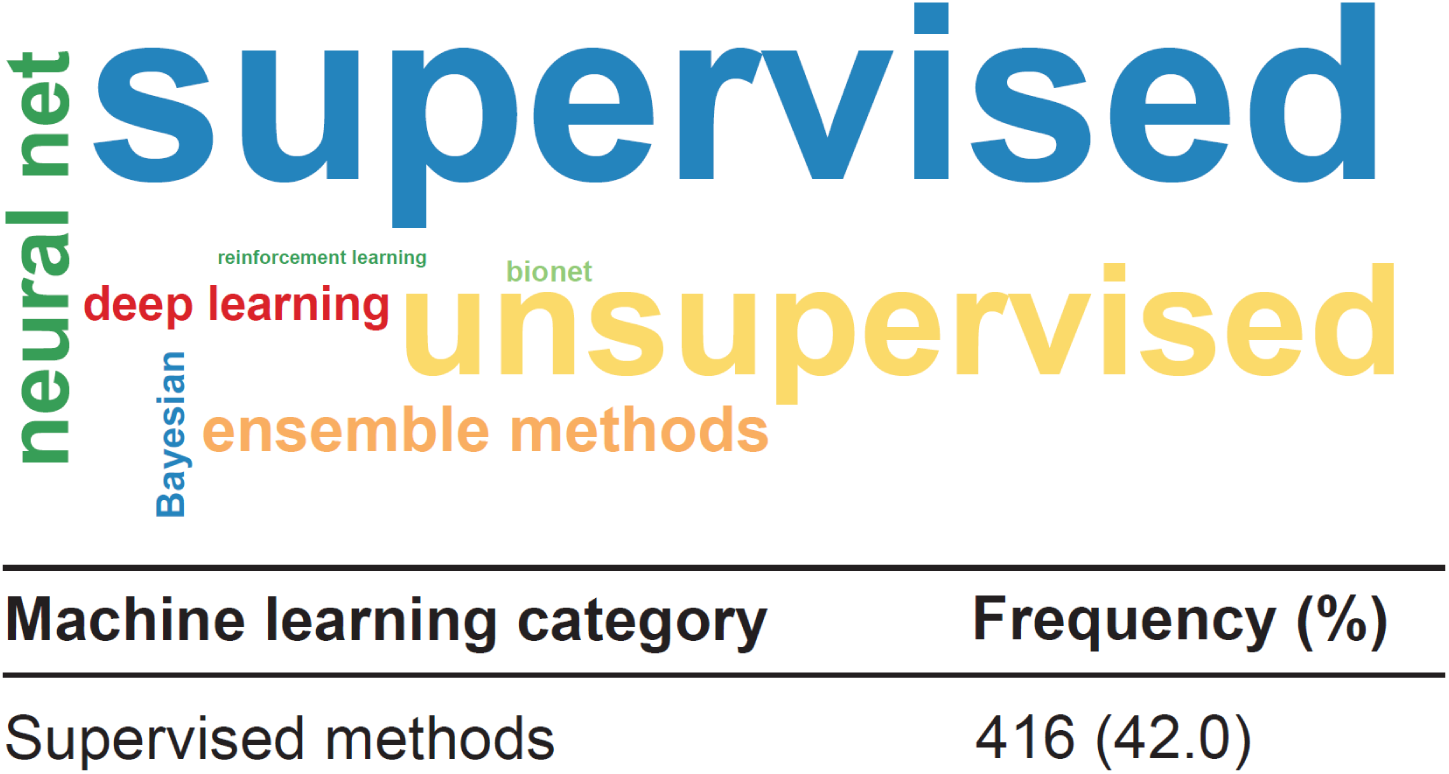
Word Cloud of Machine Learning Categories and Associated Frequency Table. *Footnote*: The size of the categories in the word cloud correspond to the frequency of the use. A machine learning method could be assigned to more than 1 theme.

## Discussion

This scoping review aimed to identify articles using ML methods in neurodegenerative disease research. Findings showed that the number of articles using ML methods in neurodegenerative disease research has increased over time. The most common data type used in ML studies was imaging data, followed by functional data. Clinical, biospecimen, genetic, electrophysiological, and molecular data were less popular. Disease diagnosis was the most common clinical aim in studies using ML methods, followed by disease prognosis and prediction of treatment effect.

Explaining the observed differences in prevalence between data types and clinical aims across diseases would require further research. We propose some avenues to explain the findings. The availability of open-source datasets may explain why some data types are used more used than others, for example, if there is more imaging data freely available than biospecimen data. For example, the Alzheimer’s Disease Neuroimaging Initiative provides access to clinical, imaging, genetic, and biomechanical biomarkers for AD for researchers.^17^ Beyond data availability, disease-specific consensus recommendations may favor the use of certain data types over others. For example, the 2021 MAGNIMS–CMSC–NAIMS consensus in MS points to the importance of magnetic resonance imaging data in diagnosing MS and supporting prognosis and treatment decisions such as therapy escalation.^18^

As an additional output of this review, we identified a list of ML methods used in the study of neurodegenerative diseases. The conduct and reporting of findings for this scoping review highlight that, although precise definitions of diseases, data types, and medical interventions were available a priori, no comparable precision was achievable for ML terms, methods, and techniques. Many “classifications” of ML techniques can be found in published papers and on websites that employ ML, but none of them have become a universally accepted standard. This is hardly surprising because the role of ML methods could be expected to vary from one field of research to another. Something that is somewhat surprising is the absence of a satisfying classification of ML in medicine, let alone in neurodegenerative diseases. Such a classification could provide more effective keywords, which, applied to our sample or future samples of studies, may lead to a deeper understanding of the state-of-the-art research in our field, and possibly indicate useful directions for further progress. A few examples of questions that should be asked to match ML methods with clinical needs are: what kind of ML approach is best indicated to (1) work with specific data types; (2) support specific medical action (e.g., treatment choice); and (3) investigate characteristics that are common to all diseases considered or to a specific disease?

Whether additional keywords would yield important information, the list of methods found in this work does not adequately reflect the current efforts of the ML community in proposing novel approaches. For instance, a central problem in ML is data integration (e.g., how to combine predictors built from different data types and/or different subjects): this has opened up several lines of investigation, such as committees or hierarchies of experts.^19–21^ Another example of an open problem in ML is seeking compromises between accuracy and interpretability (gray boxes rather than black boxes).

Our scoping review has 2 main strengths. First, it is the largest scoping review to date on the use of ML methods in neurodegenerative research. Second, it provides an additional breadth of information by including the data type and clinical aim of the studies. Our study also has limitations. First, we had to restrict the number of years included in the review to capture contemporary ML methods. Second, we limited the studies to those published in English. Third, owing to the volume of articles included in the review, we restricted our search to published articles and did not search the reference list of identified articles as well as the gray literature.

## Conclusions

The increase in application of ML techniques to understanding neurological diseases can be explained, in part, by the fact that major unmet clinical needs still exist for patients affected by these conditions, including more accurate diagnoses, disease prognostication, drug discovery, and a need for biomarkers of disease activity. Findings suggest that opportunities exist for additional research that uses ML methods, particularly for disease prognosis and prediction of treatment effect, as few studies with these clinical aims were found in our scoping review. Biospecimen and molecular data were rarely used, particularly in MS, ALS, and HD. As such, future research should focus on building databases containing these data types and on maximizing the use of these data types in neurodegenerative disease research.

## Data Availability

N/A

## Acknowledgment

The authors are grateful for Arman Alam Siddique, Shomoita Alam, Guanbo Wang, Zayd Omar, and Kaiqiong Zhao for their help with the data extraction.

## Details of Ethical Approval

Ethics approval was not required for this study as it does not include the collection of patient data. Data was retrieved from publicly available published sources.

