## Supplemental Material for "The Use of Machine Learning Methods in Neurodegenerative Disease Research: A Scoping Review"

| | |
|---|---|
| **S1 Method** | MEDLINE (PubMed) search strategy |
| **S1 Table** | Study sample size of included articles using machine learning methods in neurodegenerative diseases |
| **S2 Table** | Machine learning methods extracted from the scoping review |
| **S3 Table** | Frequency of machine learning subthemes |
| **S1 Figure** | Frequency of data type used in articles using machine learning methods for disease diagnosis, by neurodegenerative disease |
| **S2 Figure** | Proportion of data type used in articles using machine learning methods for disease diagnosis, by neurodegenerative disease |
| **S3 Figure** | Frequency of data type used in articles using machine learning methods for disease prognosis, by neurodegenerative disease |
| **S4 Figure** | Proportion of data type used in articles using machine learning methods for disease prognosis, by neurodegenerative disease |
| **S5 Figure** | Proportion of data type used in articles using machine learning methods for prediction of treatment effect, by neurodegenerative disease |
| **S6 Figure** | Proportion of data type used in articles using machine learning methods for prediction of treatment effect, by neurodegenerative disease |

**S1 Method.** MEDLINE (PubMed) search strategy

((machine learning[tiab] OR machine learning[MeSH] OR Support vector machine*[tiab] OR random forest*[tiab] OR neural network*[tiab] OR deep neural network*[tiab] OR convolutional neural network*[tiab] OR artificial neural network*[tiab] OR bayes theorem[tiab] OR k-nearest neighbor[tiab] OR k-nearest neighbour[tiab] OR gradient boost*[tiab] OR XGBoost[tiab] OR ADABoost[tiab] OR LightGBM[tiab] OR catboost[tiab] OR pattern recognition[tiab] OR pattern classification[tiab] OR transfer learning [tiab] OR supervised learning[tiab] OR unsupervised learning[tiab] OR semi-supervised learning[tiab] OR reinforcement learning[tiab] OR deep learning[tiab] OR cluster analys*[tiab] OR decision tree*[tiab] OR naive bayes*[tiab] OR k-means cluster*[tiab] OR latent Dirichlet allocation [tiab] OR discriminate analys*[tiab] OR optical character recognition*[tiab] OR chi-square automatic interaction detection*[tiab] OR computer vision[tiab] OR marker?less motion capture*[tiab] OR adaptive logic network*[tiab] OR gaussian mixture*[tiab] OR hierarchical cluster*[tiab] OR hidden markov*[tiab] OR multilayer perceptron*[tiab] OR self-organizing map*[tiab] OR mean-shift cluster*[tiab] OR density-based spatial clustering of applications with noise*[tiab] OR expectation-maximization cluster*[tiab] OR autoencoder*[tiab] OR long short-term memory[tiab] OR deep deterministic policy gradient*[tiab] OR state-action-reward-state-action*[tiab] OR deep belief network*[tiab] OR generative adversarial network*[tiab] OR restricted Boltzmann machine*[tiab] OR Skip-gram*[tiab] OR Q-learning*[tiab] OR deep Q network*[tiab] OR pose estimation*[tiab] OR Markov decision process*[tiab])
AND
(Alzheimer*[tiab] OR alzheimer's dementia[tiab] OR Alzheimer disease[mesh] OR Parkinson*[tiab] OR paralysis agitans[tiab] OR shaking palsy[tiab] OR multiple sclerosis[tiab] OR multiple sclerosis[mesh] OR amyotrophic lateral sclerosis[tiab] OR motor neuron? Disease[tiab] OR Lou Gehrig*[tiab] OR Huntington*[tiab] OR Huntington disease[mesh])
AND
("English"[Language])
AND
(diagnos*[tiab] OR detection[tiab] OR "risk of disease"[tiab] OR classif*[tiab] OR identification[tiab] OR "disease classification"[tiab] OR recognition[tiab] OR progress*[tiab] OR propagation[tiab] OR relaps*[tiab] OR prognos*[tiab] OR mortality[tiab] OR survival[tiab] OR death[tiab] OR stratification[tw] OR treatment[tw] OR therapeutic[tw] OR therap*[tw] OR patient trajector*[tw] OR treatment trajector*[tw] OR "Treatment Outcome"[MeSH] OR treatment effect[tw] OR "Precision Medicine"[MeSH] OR precision medicine[tw] OR prediction[tw] OR personalized medicine[tw] OR personalised medicine[tw] OR predictive[tw] OR individualized[tw])
AND
("1900/01/01"[Date - Publication] : "2020/12/31"[Date - Publication])
)

**S1 Table.** Study sample size of included articles using machine learning methods in neurodegenerative diseases

| | Minimum | Quarter 1 | Median | Quarter 3 | Maximum |
|---|---|---|---|---|---|
| **All articles** | 2 | 63 | 151 | 459 | 88298289 |
| **By neurodegenerative disease** | | | | | |
| Alzheimer's disease | 8 | 111 | 300 | 737 | 88298289 |
| Parkinson's disease | 2 | 40 | 80 | 195 | 229017 |
| Huntington's disease | 12 | 42 | 54 | 203 | 19369 |
| Multiple sclerosis | 5 | 48 | 94 | 232 | 8308 |
| Amyotrophic lateral sclerosis | 9 | 47 | 142 | 861 | 11908 |

**S2 Table**. Number of unique machine learning methods identified in the scoping review

| Machine learning method | Frequency |
|---|---|
| 3D attention network, 3DAN | 1 |
| 3D CNN | 1 |
| 3D densely connected convolutional networks (3D-DenseNets) | 1 |
| 3D U Net | 1 |
| AdaBoost | 25 |
| Adaptive artificial bee colony Kernel-based weighted extreme learning machine (AABC-KWELM) | 1 |
| Adaptive boosting | 1 |
| Adaptive ensemble manifold learning | 1 |
| Adaptive sparse learning | 1 |
| AlexNet Transfer Learning | 1 |
| Anatomical landmarks and directed acyclic graph (DAG) network feature learning | 1 |
| Artificial neural network | 53 |
| Autoencoder | 21 |
| Automated Symbol Digits Modalities Test | 1 |
| b-constrained-optimization-based extreme learning machine (b-COELM) | 1 |
| b-Weighted Extreme Learning Machine (b-WELM) | 1 |
| Back propagation (BP) | 1 |
| Back Propagation Neural Network | 1 |
| Back-propagation neural network of three-layer topology | 1 |
| Bagged decision tree | 3 |
| Bagging | 1 |
| Bagging ensemble algorithm | 2 |
| Bagging-based ensemble regression | 1 |
| Bayesian adaptive regression trees | 1 |
| Bayesian Belief Networks | 1 |
| Bayesian Lasso | 1 |
| Bayesian Machine Learning | 1 |
| Bayesian Multitask with Structure Learning | 1 |
| Bayesian network | 4 |
| Bayesian Neural Network | 1 |
| Bayesian Ridge Regression | 1 |
| BayesNet | 1 |
| Behavior score-embedded encoder network (BSEN) | 1 |
| Bidirectional long short-term memory (BiLSTM) | 3 |
| Biorthogonal wavelet transform | 1 |
| Boosted decision tree | 2 |
| Boosted Logistic regression | 1 |
| Bootstrap Stage-Wise Model Selection (BSWiMS) | 1 |
| Canonical correlation analysis | 2 |
| Cascade ensemble | 1 |
| Cascaded multi-column RVFL+ (cmcRVFL+) framework | 1 |
| Classification and Regression Tree (CART) | 5 |
| Cluster Analysis | 2 |
| Clustered Multi-Task Learning (CMTL) | 1 |
| CNN-Stochastic Coordinate Coding (MSCC) | 1 |
| Combination of L2;pnorm of prediction loss function and L2;qnorm of feature selection | 1 |
| Convolution Encoder Network | 1 |
| Convolutional neural network | 166 |
| Coupled boosting (CB) | 1 |
| CoxBoost | 1 |
| Cubist | 1 |
| Decision tree | 126 |

| | |
|---|---|
| Decorrelated neural network ensembles | 1 |
| Deep 3D convolutional encoder networks | 1 |
| Deep belief network | 3 |
| Deep Boltzmann Machine | 1 |
| Deep CNN | 1 |
| Deep Ensemble Learning | 1 |
| Deep Ensemble Sparse Regression Network (DeepESRNet) | 1 |
| Deep learning | 29 |
| Deep multi-modal fusion network | 1 |
| Deep network-based feature selection (DN-FS) | 1 |
| Deep neural mapping large margin distribution machine (DNMLDM) | 1 |
| Deep neural network | 42 |
| Deep Polynomial Networks Algorithm | 1 |
| Deep residual neural network (ResNet) | 1 |
| Deep weighted S2MTL (DW-S2MTL) | 1 |
| Density-based clustering (Den-sity-Based Spatial Clustering of Applications with Noise [DBSCAN]) | 1 |
| Discriminant Dictionary Learning | 1 |
| Discriminate analysis | 85 |
| Discriminative contractive slab and spike convolutional restricted Boltzmann Machine (DCssCRBM) | 1 |
| Dynamic Tree Cut algorithm | 1 |
| Echo state networks (ESNs) | 1 |
| Elastic Net | 23 |
| Elastic Net Linear Regression | 1 |
| Elastic net regression | 1 |
| Elastic net regularized logistic regression (EN-RLR) | 4 |
| Elastic net regularized regression model | 1 |
| Empirical Bayes | 1 |
| Empirical Bayes Transfer Learning | 1 |
| Enhanced Probabilistic Neural Network | 1 |
| Ensemble classification | 2 |
| Ensemble learning | 1 |
| Ensemble Linear Discriminant (ELD) | 1 |
| Ensemble method | 2 |
| Ensemble neural networks based on Akaike information criterion (ENN-AIC) | 1 |
| Ensemble transfer learning (ETL) | 1 |
| Ensemble-bagged trees | 1 |
| Ensemble-subspace discriminant | 1 |
| Epinet | 1 |
| Evolutionary Wavelet Neural Networks | 1 |
| Evolutionary weighted random support vector machine cluster (EWRSVMC) | 1 |
| Expectation Maximization Diverse Density (EM-DD) | 1 |
| Expectation-maximization cluster | 2 |
| Extra Trees Classifier | 1 |
| Extreme gradient boosting | 1 |
| Extreme Gradient-Boosted Decision Trees (XGB) | 1 |
| Extreme Learning Machine (ELM) | 15 |
| F neuro-fuzzy classifiers | 1 |
| Factor Analysis | 1 |
| Fast-multiple kernel learning framework | 1 |
| FC Net | 1 |
| FCM based Weighted Probabilistic Neural Network (FWPNN) | 1 |
| Feed Forward Neural Networks | 1 |
| Forward–Backward Support Vector Machine | 1 |
| Fully bayesian longitudinal unsupervised learning | 1 |
| Fully stacked bidirectional long short-term memory (FSBi-LSTM) | 1 |
| Functional logistic regression | 1 |

| | |
|---|---|
| Fused group lasso | 2 |
| Fuzzy C-mean | 1 |
| Fuzzy Granular Hyperplane Classifiers | 1 |
| Fuzzy Rough Classifier | 1 |
| Gaussian discriminative component analysis | 1 |
| Gaussian Kernel | 1 |
| Gaussian mixture | 11 |
| Gaussian Naïve Bayes | 1 |
| Gaussian Process Classifier | 1 |
| Gaussian process regression | 2 |
| General additive (GAM) | 1 |
| Generalized Boosting Model | 1 |
| Generalized composite multiscale entropy vector | 1 |
| Generalized logistic regression analysis | 1 |
| Generalized matrix learning vector quantization | 1 |
| Generalized Multiple Kernel Learning | 1 |
| Generative adversarial network | 4 |
| GLMBoost | 1 |
| Gradient boost | 59 |
| Graph based Transductive learning (GTL) | 1 |
| Graph convolution network | 3 |
| Graph Filtration | 1 |
| Graph Net | 2 |
| Graph-CNN | 1 |
| Graph-guided multi-task learning | 2 |
| Group independent component analysis | 1 |
| Group Lasso | 7 |
| Hidden markov | 24 |
| Hierarchical Ascendant Classification (HAC) | 1 |
| Hierarchical Classification | 1 |
| Hierarchical cluster | 16 |
| Hierarchical cluster analysis | 1 |
| Hierarchical CNN | 1 |
| Hierarchical Graph Neural Network (hi-GCN) | 1 |
| High-Order Graph Matching method | 1 |
| Import Vector Machine | 1 |
| Independent component analysis | 2 |
| Inductive Fuzzy Classifier | 1 |
| Instance-based Learning (IBL) | 1 |
| Iterative Discriminative Axis Parallel Rectangle (ID-APR) | 1 |
| J48 classifier | 1 |
| Joint Coupled-Feature Representation and Coupled Boosting method | 1 |
| Joint feature-sample selection (JFSS) | 1 |
| Joint Linear and Logistic Regression with 2, 1-norm regularization (JLLR) | 1 |
| k-means cluster | 27 |
| k-nearest neighbor | 159 |
| Kernel Principal Component Analysis | 1 |
| Kernel Ridge Regression (KRR) | 1 |
| Kernel-based extreme learning machine (KELM) | 1 |
| Kohonen map | 1 |
| L1-norm regularized logistic regression | 1 |
| L2-norm regularized logistic regression | 1 |
| Large Memory STorage And Retrieval (LAMSTAR) neural network, | 1 |
| LASSO | 31 |
| LASSO linear regression | 1 |
| Lasso logistic regression | 40 |

| | |
|---|---|
| LASSO regularized Cox regression model | 1 |
| Latent class mixed modelling (LCMM) | 1 |
| Latent Profile Analysis (LPA) | 1 |
| Latent space representation | 1 |
| Layer-wise relevance propagation (LRP | 1 |
| Least-Square Sparse Regression Elastic Net | 1 |
| LightGBM | 1 |
| Linear discriminant analysis | 4 |
| Linear Mixed Effects | 1 |
| Linear Regression | 4 |
| Linked independent component analysis | 1 |
| Local naïve Bayes brain network model | 1 |
| Locality Preserving Projections (LPP) | 1 |
| Locally Linear Embedding (LLE) | 1 |
| LOCALLY WEIGHTED PENALIZED REGRESSION | 1 |
| Logistic regression | 161 |
| Logistic regression with Elastic Net regularization | 1 |
| Logistic Sparse Regression Elastic Net | 1 |
| Logitboost | 2 |
| LOLIMOT (Local Linear Model Trees) | 2 |
| Long short term memory neural networks | 8 |
| Long Short-Term Memory (LSTM) | 2 |
| Longitudinal Siamese network (LSN) | 1 |
| M5-model trees | 1 |
| MANCOVA | 1 |
| Manifold learning | 1 |
| Manifold regularized Multi-Task Feature Selection | 1 |
| Mean Multi-Kernel SVM | 1 |
| Missingness-Aware Temporal Convolutional Hitting-time Network | 1 |
| Mixed effect linear model | 1 |
| Mixed effects model | 1 |
| Multi Kernel Learning | 2 |
| Multi Modal Multi Task | 1 |
| Multi Output Linear Regression with 2, 1-norm regularization (MOLR) | 1 |
| Multi Task Learning | 1 |
| Multi-feature Kernel SCDDL (MKSCDDL) | 1 |
| Multi-Hypergraph based Classification | 1 |
| Multi-kernel method with LASSO feature selection | 1 |
| Multi-Kernel SVM | 4 |
| Multi-modal feature selection method | 1 |
| Multi-Modal Multi-Task | 5 |
| Multi-Task Deep Learning (MTDL) | 1 |
| Multi-task Stochastic Coordinate Coding | 1 |
| Multi-layer clustering | 1 |
| MultiBoost-AB and J48 | 1 |
| MultiBoost-AB and Simple Cart | 1 |
| Multifactor dimensionality reduction (MDR) | 1 |
| Multilayer perceptron | 41 |
| Multimodal random forest | 1 |
| Multinomial regression | 1 |
| Multiple Instance Neural Network (MI-NN) | 1 |
| Multiple Instance Support Vector Machine (MISVM) | 1 |
| Multiple kernel learning (MKL) | 6 |
| Multiple linear regression | 1 |
| Multiple-output elastic-net regression | 1 |
| Multivariate linear models | 1 |

| | |
|---|---|
| Naïve Bayes | 85 |
| Naive Support Vector Machine (Naive-SVM) | 1 |
| Nearest Shrunken Centroid | 1 |
| Neighborhood Representation Local Binary Patterns method | 1 |
| Network Diffusion Model | 1 |
| Network wide association method | 1 |
| Neural network | 66 |
| Nu-Support Vector Classification | 1 |
| Online sequential extreme learning machine (OS-ELM) | 1 |
| Optimal Deep Learning | 1 |
| Optimum-Path Forest (OPF) | 1 |
| Ordinary Least Squares (OLS) | 1 |
| Orthogonal least squares algorithm | 1 |
| Overlapping group lasso | 1 |
| Partial least square discriminant analysis | 3 |
| Partial least squares | 3 |
| Partitioning around medoids (PAM) | 1 |
| Passive Aggressive Regression | 1 |
| Pearson correlation coefficient-based feature selection | 1 |
| Positive Transfer Learning | 1 |
| Principal component analyses (PCA) | 29 |
| Principal Component Regression | 1 |
| Principal-component-based logistic regression (PC-LR) | 1 |
| Probabilistic neural network | 1 |
| Q-learning | 4 |
| Radial Basis Function Network | 3 |
| Radial Basis kernels (SVM-R) | 1 |
| Random forest | 310 |
| Random forest -MaxStat | 1 |
| Random forest -survival | 3 |
| Random Linear Oracle | 1 |
| Random Multimodal Deep Learning (RMDL) | 1 |
| Random neural network cluster | 1 |
| Random Undersampling boosting | 1 |
| RandomCommittee and ExtraTree | 1 |
| Recurrent neural network | 28 |
| Recursive Partitioning and Regression Trees (RPART) | 1 |
| Recursive-clustering | 1 |
| Regression | 1 |
| Regression forest | 1 |
| Regularized Bagged - Canonical Correlation Analysis (RB-CCA) | 1 |
| Regularized Elastic Net Logistic Regression | 1 |
| Regularized logistic regression | 1 |
| Reinforcement Learning | 7 |
| Relevance Vector Machine | 1 |
| Relevance vector regression (RVR) | 1 |
| Repeated Incremental Pruning to Produce Error Reduction (RIPPER) | 2 |
| Residual Neural Networks | 1 |
| ResNet | 1 |
| Resource-efficient oblique tree with power-efficient regularization (ResOT-PE) | 1 |
| Restricted Boltzmann machine | 4 |
| Ridge Classifier | 1 |
| Ridge logistic regression | 6 |
| Ridge regression | 12 |
| Robust multi-label transfer feature learning (rMLTFL) | 2 |
| Safe Semi Supervised SVM | 1 |

| | |
|---|---|
| Self-paced learning (SPL) | 1 |
| Semi-supervised Multi-view learning Clustering architecture technology (SMC) | 1 |
| Semi-unsupervised 3C strategy (Categorize, Cluster, Classify—CCC) | 1 |
| Soft max | 1 |
| Sparse and functional principal component analysis (SFPCA) | 1 |
| Sparse high-order interaction model with rejection option (SHIMR) | 1 |
| Sparse Joint Classification and Regression | 2 |
| Sparse multi-task learning | 1 |
| Sparse regression | 1 |
| Sparse-response deep belief network (SR-DBN) model | 1 |
| Sparsity Preserving Projections (SPP) | 1 |
| Spatial-Temporal Convolutional-Recurrent Neural Network | 1 |
| Stacked Classifier | 1 |
| Stacked long short-term memory (SLSTM) | 1 |
| SubMito-XGBoost | 1 |
| Supervised Cluster Analysis | 1 |
| Supervised ensemble learning | 1 |
| Supervised Euclidean distance-based clustering method | 1 |
| Supervised Joint Classification and Regression (JCR) | 1 |
| Supervised within-Class-Similar Discriminative Dictionary Learning | 1 |
| Support vector data description | 1 |
| Support vector machine | 651 |
| Support vector machine (well SVM) | 1 |
| Support vector machine +RBF kernel | 1 |
| Support vector machine with gaussian kernels | 1 |
| Support vector machine-based recursive feature elimination (SVM-RFE) | 1 |
| Support Vector Regression | 1 |
| Swarm | 1 |
| Symbolic Regression | 1 |
| Temporal Cluster Analysis | 1 |
| Transfer learning | 3 |
| Tree guided sparse learning | 1 |
| Tree regression | 1 |
| Two-step clustering | 1 |
| Universum support vector machine based recursive feature elimination (USVM-RFE) | 1 |
| Unsupervised Binary Trees | 1 |
| Unsupervised Deep Autoencoder | 1 |
| Unsupervised Diffusion Component Analysis | 1 |
| Unsupervised learning | 1 |
| Unsupervised machine learning model | 1 |
| VGG16 | 1 |
| View-aligned hypergraph classification | 1 |
| Wavelet Scattering Transform with Support Vector Machine (WST-SVM) | 1 |
| Weighted gene co-expression network analysis | 1 |
| Weighted visibility graph | 1 |
| XGBoost | 5 |
| XGBoost-Linear | 1 |
| XGBoost-Tree | 1 |
| **Total** | **2,734** |

**S3 Table.** Frequency of machine learning subthemes

| Theme | Machine learning subtheme | Frequency [a] |
|---|---|---|
| Supervised methods | Supervised | 125 |
| Supervised methods | Classification | 72 |
| Supervised methods | Discrimination | 11 |
| Supervised methods | Support vector machine | 19 |
| Supervised methods | Naïve Bayes | 3 |
| Supervised methods | Regression | 64 |
| Supervised methods | Multiple response | 7 |
| Supervised methods | Longitudinal response | 7 |
| Supervised methods | Linear mixed model | 4 |
| Supervised methods | Generalized regression | 19 |
| Supervised methods | Gaussian process | 2 |
| Supervised methods | survival response | 3 |
| Supervised methods | penalized | 28 |
| Supervised methods | LASSO and generalizations | 8 |
| Supervised methods | Elastic net and generalization | 10 |
| Supervised methods | Decision tree/CART | 26 |
| Supervised methods | Random forest | 6 |
| Supervised methods | Functional data | 2 |
| Unsupervised methods | Semi-supervised | 8 |
| Unsupervised methods | Unsupervised | 46 |
| Unsupervised methods | Clustering | 23 |
| Unsupervised methods | Hierarchical | 6 |
| Unsupervised methods | k-nearest neighbors | 2 |
| Unsupervised methods | Spectral clustering | 0 |
| Unsupervised methods | Association models | 6 |
| Unsupervised methods | Model-based | 64 |
| Unsupervised methods | Case-based | 17 |
| Unsupervised methods | Instance-based | 2 |
| Unsupervised methods | Dimensionality reduction | 20 |
| Unsupervised methods | Latent space | 4 |
| Unsupervised methods | Feature selection | 13 |
| Unsupervised methods | Sample selection | 1 |
| Unsupervised methods | Kernel methods | 15 |
| Unsupervised methods | Manifold methods | 4 |
| Bayesian | Bayesian approach | 15 |
| Bayesian | Bayesian/Belief network | 5 |
| Ensemble methods | Ensemble methods | 49 |

| | | |
|---|---|---|
| Ensemble methods | Boosting | 17 |
| Ensemble methods | Baggins | 6 |
| Ensemble methods | ADAboost | 1 |
| Ensemble methods | X gradient boosting | 5 |
| Reinforcement learning | Reinforcement learning | 3 |
| Neural net | Deep learning | 90 |
| Neural net | Artificial neural network | 40 |
| Neural net | Extreme learning machine | 8 |
| Neural net | Convolutional neural network | 17 |
| Neural net | Recurrent neural network | 7 |
| Neural net | Based on fuzzy logic | 8 |
| Neural net | Transfer learning | 6 |
| Deep learning | Multi-task | 12 |
| Deep learning | Multi-mode | 9 |
| Deep learning | Multiple instances | 3 |
| Deep learning | Optimization | 7 |
| Deep learning | Algorithm | 17 |
| Deep learning | Method involving graph | 14 |
| Bionet | BioNet | 6 |

[a] A machine learning method could be assigned to more than one subtheme

**S1 Fig**. Frequency of data type used in articles using machine learning methods for disease diagnosis, by neurodegenerative disease [a]

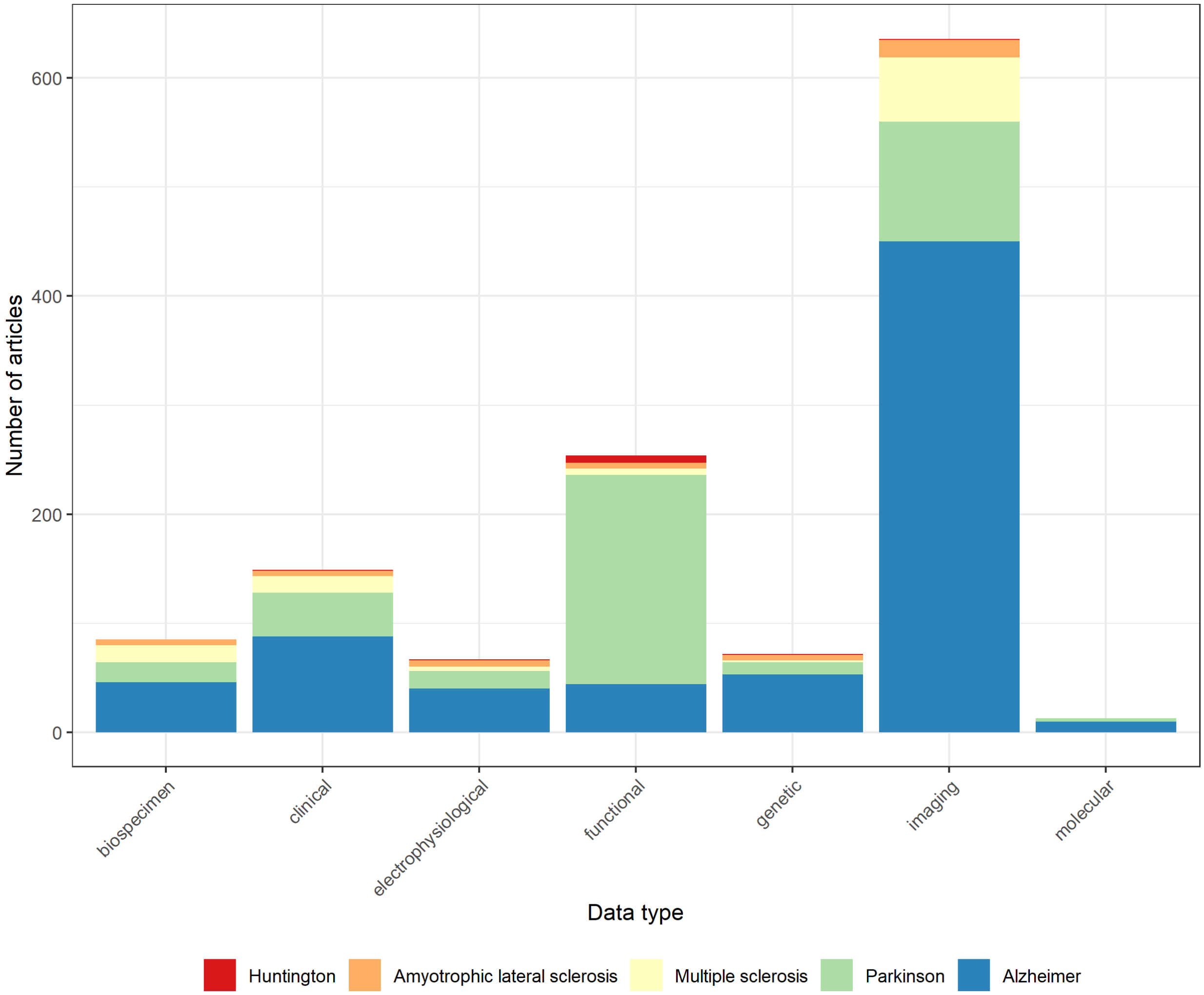


[a] An article can include more than one data type

**S2 Fig**. Proportion of data type used in articles using machine learning methods for disease diagnosis, by neurodegenerative disease

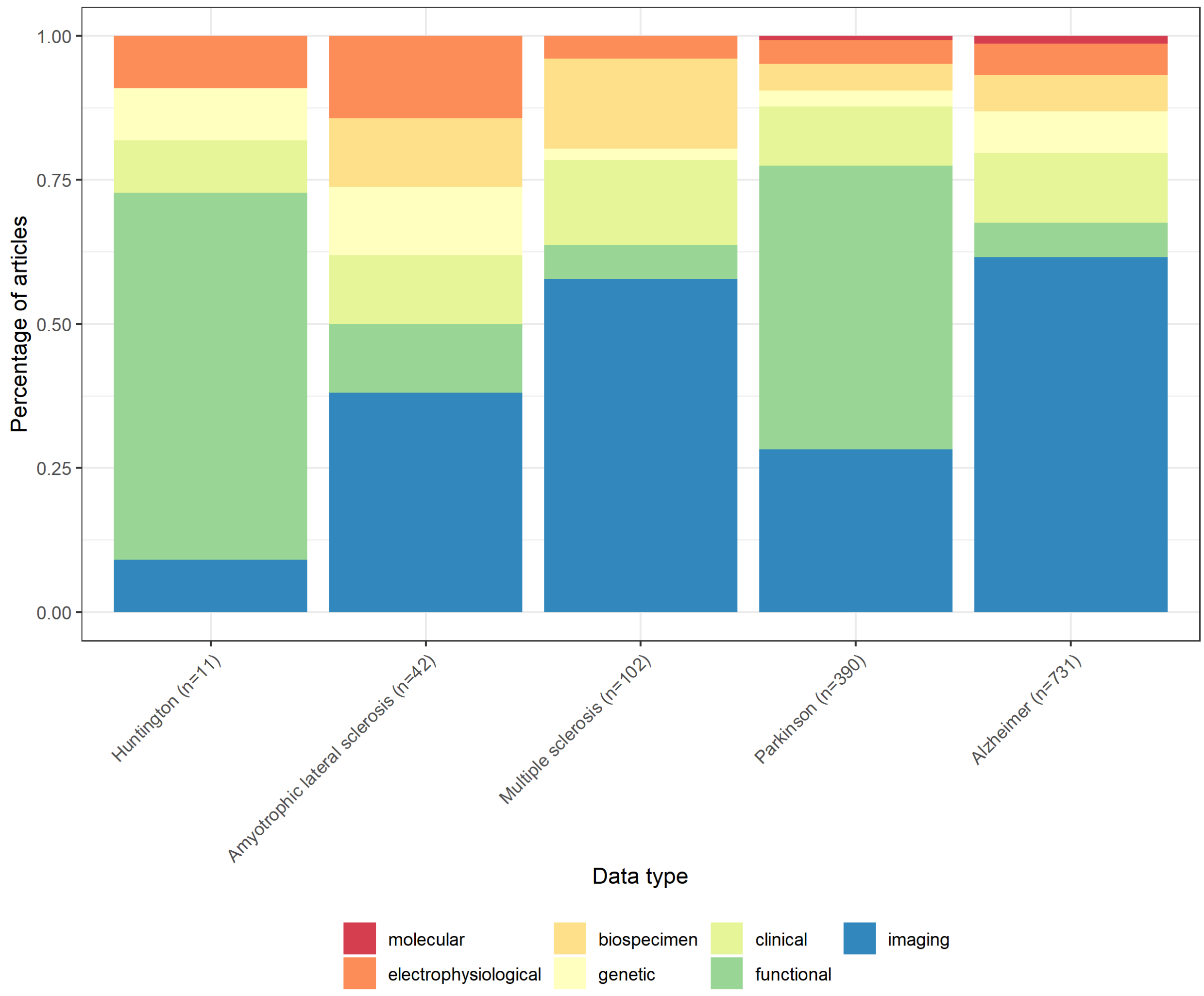

**S3 Fig**. Frequency of data type used in articles using machine learning methods for disease prognosis, by neurodegenerative disease [a]

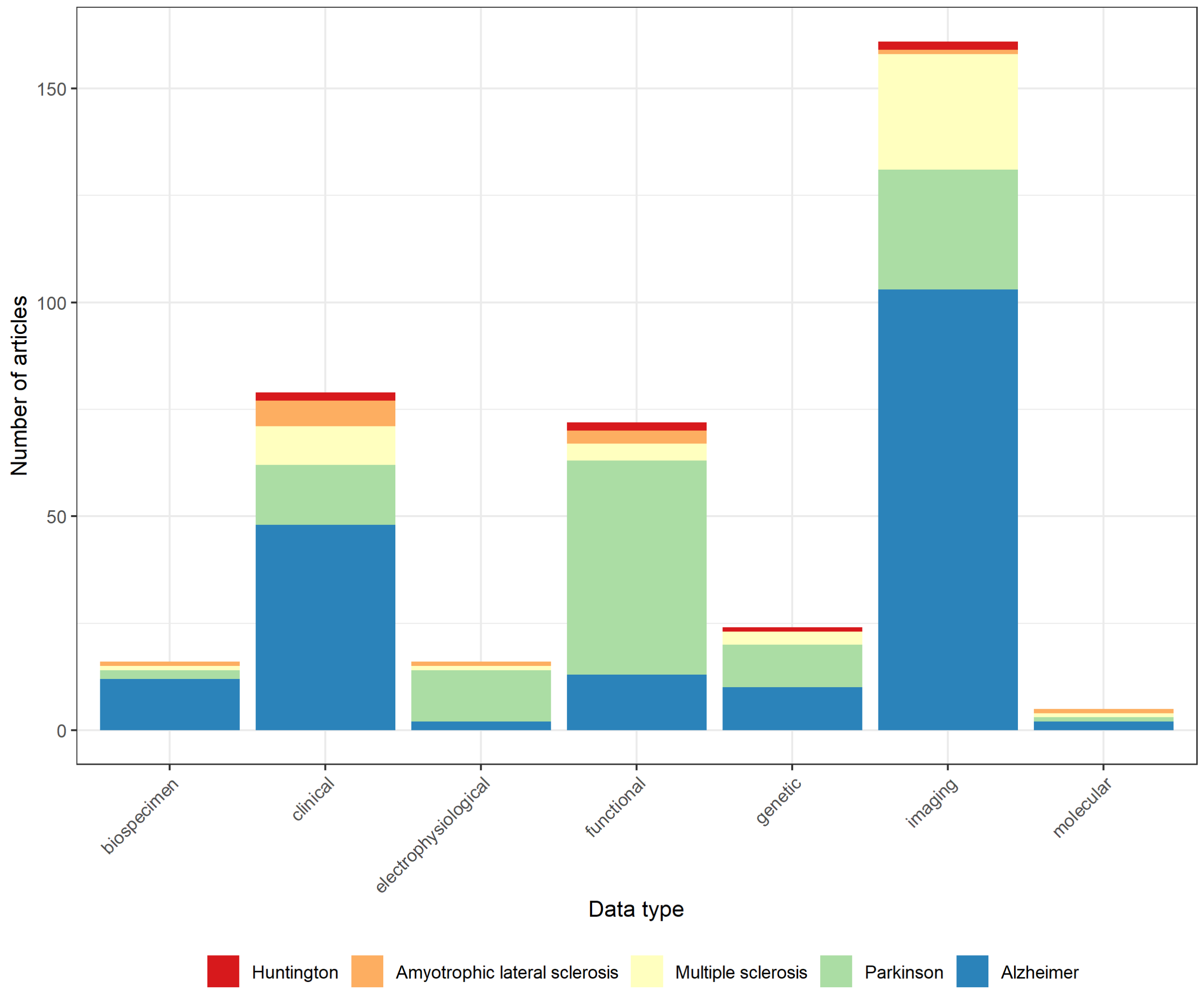


[a] An article can include more than one data type

**S4 Fig**. Proportion of data type used in articles using machine learning methods for disease prognosis, by neurodegenerative disease

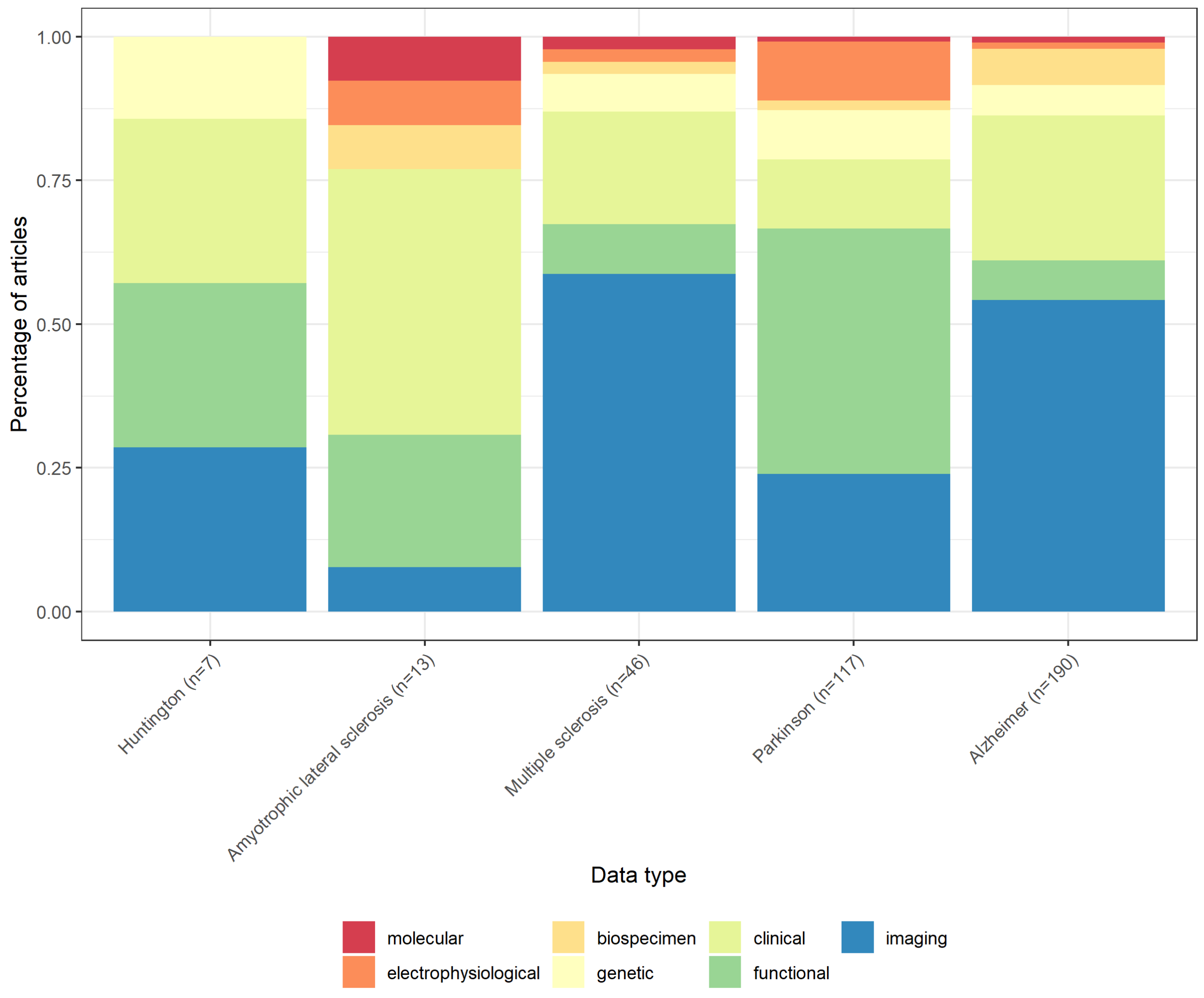

**S5 Fig**. Frequency of data type used in articles using machine learning methods for prediction of treatment effect, by neurodegenerative disease [a]

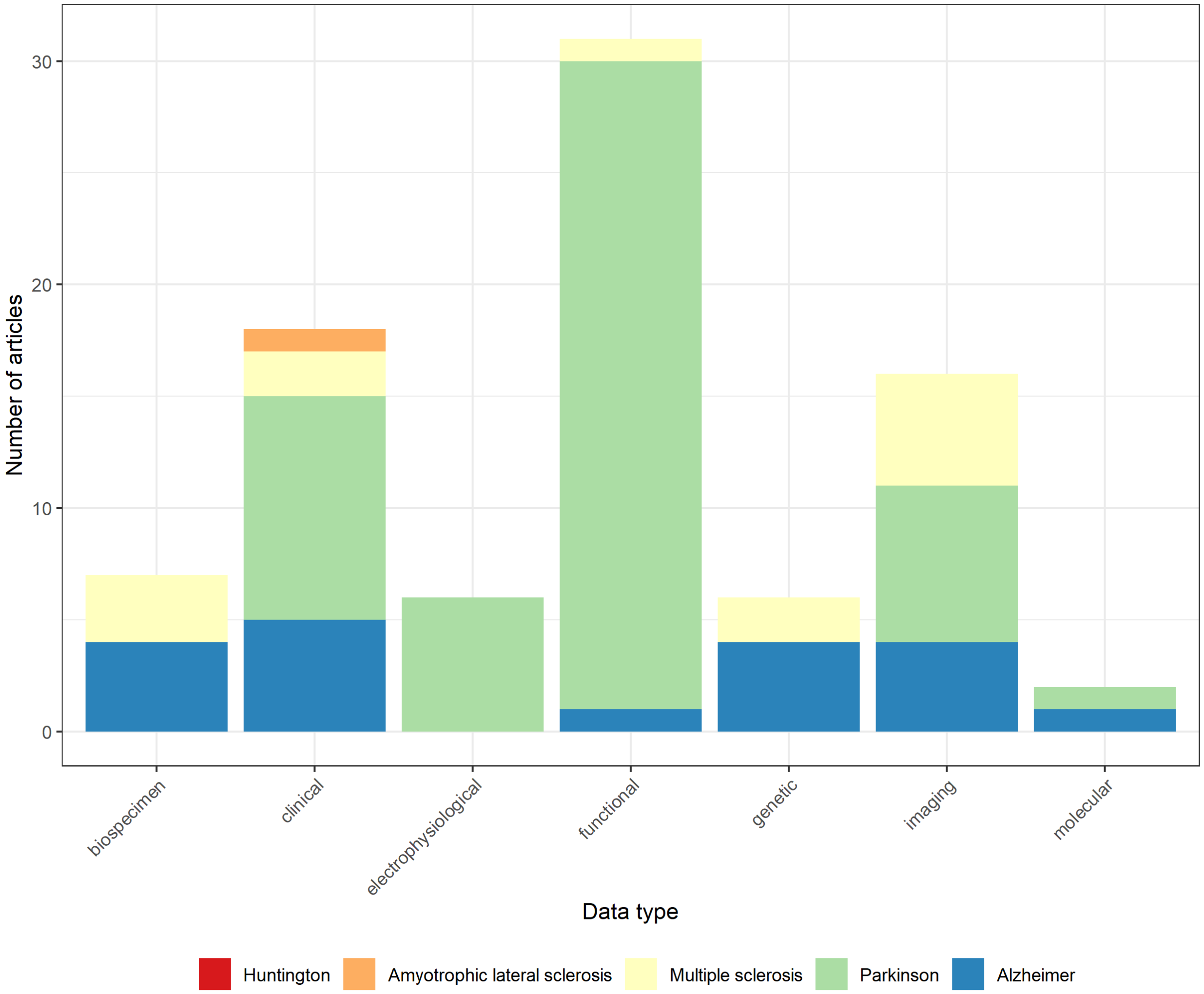


[a] An article can include more than one data type

**S6 Fig**. Proportion of data type used in articles using machine learning methods for prediction of treatment effect, by neurodegenerative disease

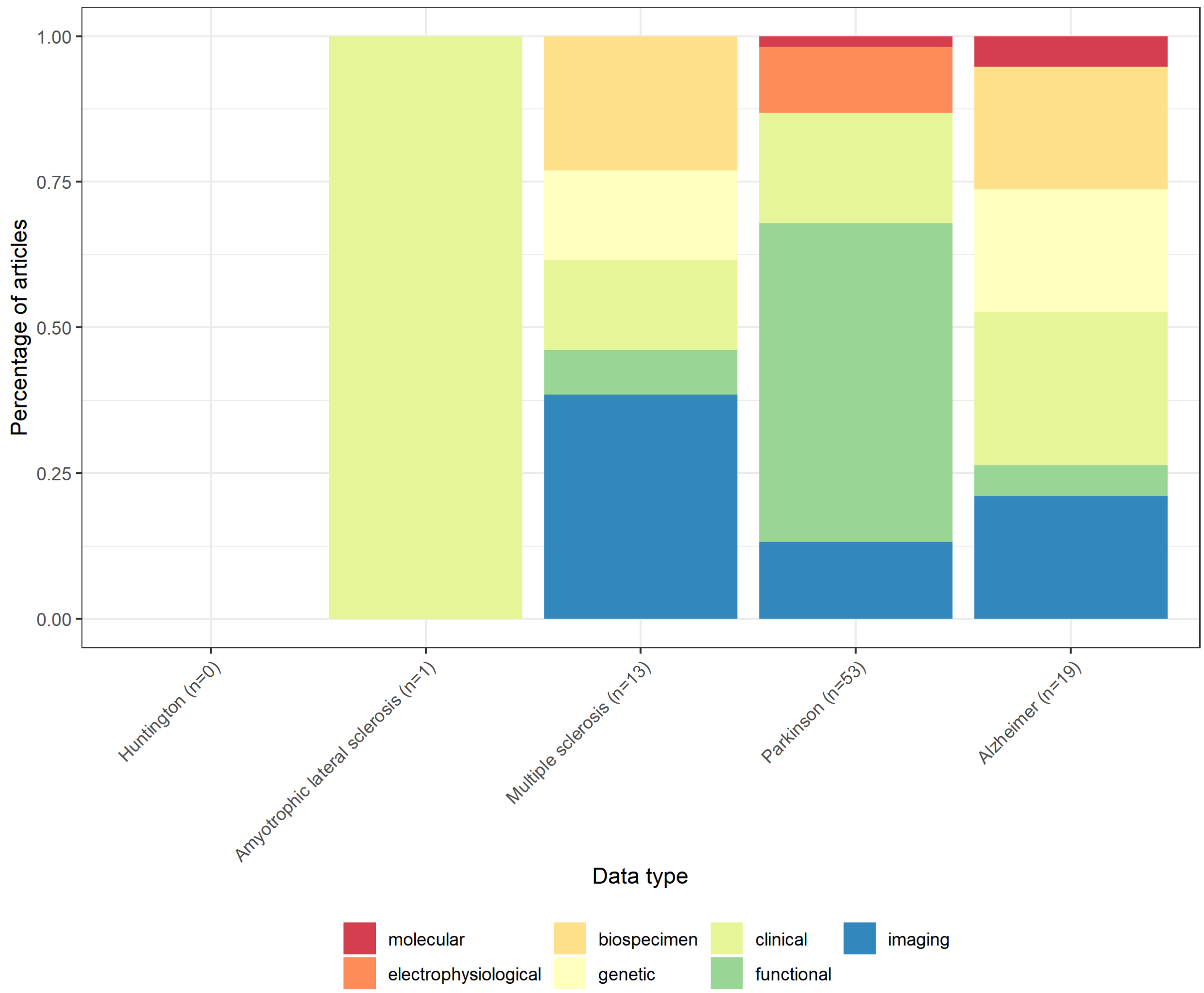
